# Sun, Sleep, and Satisfaction: Mediating Role of Depression and Source of Endogeneity among Middle-aged and Older Adults in China

**DOI:** 10.1101/2022.01.09.22268931

**Authors:** Xiao Han, Jun Li

**Author notes:** (Corresponding Author) Postal Address: Peking University Health Science Center, 38 Xueyuan Rd, Haidian, District, Beijing, China, 100191. Postal Address: Peking University Health Science Center, 38 Xueyuan Road, Haidian, District, Beijing, China, 100191.

## Abstract

**Purpose:** To examine: (i) depression as a mediator in effects of sleep duration and quality on life satisfaction (LS), (ii) source of endogeneity in self-reported data on sleep, and (iii) predictive power of sleep duration and quality on LS.

**Methods:** Panel data of 22,674 observations from the China Health and Retirement Longitudinal Survey (2015 & 2018) was used. Sleep was assessed with self-reported duration and quality. Depression was measured by the 10-question version of the Center for Epidemiological Survey - Depression. LS was rated by five scales. Fixed-effects ordered logit models were used to determine the effect of sleep duration and quality on life satisfaction and the mediating role of depression. We used instrumental variable strategy to explore the source of endogeneity. Information value and random forest model were used to examine the predictive power of sleep measures duration and quality.

**Results:** Sleep duration and quality were found to improve life satisfaction via lower depression score. Non-agricultural employed population with urban *hukou* (household registration) accounted for the endogeneity, but the instrument variable sunset failed the weak instrument test. Sleep measures were found to predict life satisfaction, especially for the lower life satisfaction groups.

**Conclusion:** Our findings suggest the importance of sleep and the study of the associations between solar cues, social schedules, and sleep. Policy makers of social care of older adults might consider sleep intervention among this population.

## Introduction

Subjective well-being (SWB) is an important measure of quality of life. Compared with traditional economic indicators (for example, income, employment), it can better reflect subjective quality of life and is sensitive to social conditions, and thus increasingly favored by researchers (Diener & Tay, 2015).

The cognitive dimension of SWB, life satisfaction (LS), represents an individual’s global evaluation of life (Diener et al., 1985). LS itself is one of the pursuits of a happier life. Higher LS is positively associated with life events such as marriage and child birth, and negatively associated with job disruptions and divorce (Luhmann et al., 2012). In Chinese adults, LS reduces mortality risk (Xu et al., 2021), mediates the protective effect of security on social trust (Shao et al., 2021), and is the only dimension of SWB that mediating the positive effect of social cohesion on physical health by improving it (Zhang et al., 2021).

Sleep proves to be one of the effective interventions of SWB (Diener & Biswas-Diener, 2019; Weinberg et al., 2016). Short sleep duration and poor sleep quality are inversely related with life satisfaction, partially mediated by depression in elderly Chinese (Zhi et al., 2016). Sleep quality and variability of sleep duration, instead of sleep duration, is strongly associated with life satisfaction in Norwegian university students (Ness & Saksvik-Lehouillier, 2018). Moreover, growing prevalence of sleep deprivation may pose a threat to population health and bring significant burden to the healthcare system (Roenneberg, 2013). Therefore, it is meaningful to examine effects of sleep duration/quality on LS in middle-aged and older adults.

On the other hand, the mediating role of depression might be explained by negative associations with sleep and LS. Depression can increase the risk of frequent sleep disruption (Fang et al., 2019; Perlis et al., 1997) and may be correlated with compromised quality of life (Duong, 2021; Kim & Ko, 2018) and neuro-psychological functions (Sha et al., 2019) among older adults.

However, these findings are mostly based on subjects in Europe and North America (Diener et al., 2018) and may not be applicable to people in Asia. There is regional heterogeneity in associations between socioeconomic indicators and LS, sleep duration and LS. For example, one study in Asia found that the effects of gender and age on LS are insignificant except in Central & West Asia (Ngoo et al., 2021). The best sleep duration was found to be around 8 hours (Piper, 2016) in Germans for maximal LS, but the optimal sleep duration for LS might differ in Asian populations (Diener et al., 2018).

Sleep is usually self-reported in social surveys (Diener et al., 2018). Such data, in contrast to objective sleep in a daily setting, have different psychological and biological correlates that could lead to inconsistent conclusions about sleep and well-being (Jackowska et al., 2016). Objectively-measured sleep is related with executive functioning broadly and self-reported sleep with conceptual flexibility in particular (Bernstein et al., 2019). Fixed-effects models may help eliminate the self-reported bias, but determining source of endogeneity requires instrumental variable, an econometric model. Sunset time has proved to be a candidate instrumental variable relevant among urban non-agricultural employed population (Giuntella et al., 2017).

Previous studies, if not cross-sectional, mostly did not control for individual fixed effects and self-reported bias that are detrimental for the elucidation of the causality between sleep and LS. Therefore, we conducted this fixed-effects longitudinal analysis and used objectively measured data, sunset time, as the instrument variable.

Using the nationally representative data on sleep and LS of middle-aged and older adults and the sunset time of the subjects’ geographical location in China, this study aims to explore the effects of sleep duration/quality on life satisfaction, the possible mediating effect of depression, and the predictive power of sleep measures for LS.

Based on the literature, we proposed the following hypotheses:

**H1** Duration and quality of sleep have a positive effect on LS.

**H2** Depression mediates the effect of sleep on LS. Specifically, depression is negatively associated with sleep duration/quality and LS.

**H3** Urban non-agricultural employed population accounts for endogeneity.

**H4** Duration and quality of sleep predict LS, especially in identifying poor LS.

## Methods

### Data

Data from the third (2015) and fourth (2018) waves of the China Health and Retirement Longitudinal Survey (CHARLS) were used. CHARLS collected the data of demographics, economic and health status, family conditions, and social conditions at the community/city level of the participants. The baseline survey was conducted in 450 communities from 150 counties/districts in 28 provinces. For details of the survey, see the Survey website (http://charls.pku.edu.cn/) and relevant literature (Zhao et al., 2014; Zhao et al., 2020; Zhao et al., 2013). After deleting 1,273 observations with missing or extreme values of sleep duration in the two waves (37,620 observations in total), 7,376 observations for subjects aged outside of the 45-to-70-years range, 151 observations from Xinjiang (where a time different from other areas in China was used) and 6,146 observations inapplicable (due to incompleteness of data) to longitudinal analysis, 22,674 observations remained (for details of subjects, see Table 1 and 2).

**Table 1.** Descriptive statistics of continuous variables.

|  | 2015 |  |  | 2018 |  |  |
| --- | --- | --- | --- | --- | --- | --- |
|  | Mean | SD | N | Mean | SD | N |
| Life Satisfaction | 3.39 | 0.77 | 11,337 | 3.24 | 0.79 | 11,337 |
| Sleep Hours | 6.42 | 1.72 | 11,337 | 6.21 | 1.74 | 11,337 |
| Napping Time | 38.10 | 44.04 | 11,337 | 38.66 | 43.75 | 11,337 |
| Sleep Quality | 2.02 | 1.19 | 11,312 | 1.88 | 1.21 | 11,224 |
| Schooling Years | 5.76 | 4.21 | 11,337 | 5.64 | 4.17 | 11,337 |
| PCE (logarithm) | 9.73 | 0.94 | 11,337 | 10.03 | 0.91 | 11,326 |
| Age | 55.84 | 6.51 | 11,337 | 58.72 | 6.51 | 11,337 |
| CESD | 7.57 | 6.18 | 11,337 | 8.28 | 6.39 | 11,337 |
| GDP (logarithm) | 5.26 | 0.89 | 11,337 | 5.44 | 0.92 | 11,337 |
| Population (logarithm) | 8.51 | 0.58 | 11,337 | 8.52 | 0.59 | 11,337 |

Note: SD = standard deviation; PCE = per capita expenditure at household level; CESD = Center for Epidemiological Survey - Depression; GDP = gross domestic product at city level

**Table 2.** Descriptive statistics of categorical variables.

|  | 2015 |  | 2018 |  |
| --- | --- | --- | --- | --- |
|  | % | N | % | N |
| Urban *Hukou* | 19.3 | 2,191 | 20.3 | 2,301 |
| Female | 51.8 | 5,870 | 51.7 | 5,862 |
| Married | 92.5 | 10,483 | 90.6 | 10,269 |
| Social Participation | 57.3 | 6,499 | 55.9 | 6,338 |
| Physical Disability | 23.3 | 2,555 | 23.9 | 2,696 |
| Employment |  |  |  |  |
| Non-agricultural Employed | 28.1 | 3,122 | 23.6 | 2,658 |
| Self-employed | 12.5 | 1,394 | 9.2 | 1,031 |
| Farmer | 40.0 | 4,446 | 38.9 | 4,373 |
| No work | 19.3 | 2,148 | 28.3 | 3,185 |
| Terrain |  |  |  |  |
| Plain | 40.9 | 4,627 | 40.9 | 4,627 |
| Hills | 30.4 | 3,434 | 30.4 | 3,434 |
| Mountainous Region | 21.0 | 2,379 | 21.0 | 2,379 |
| Plateau | 4.5 | 514 | 4.5 | 514 |
| Basin | 3.1 | 351 | 3.1 | 351 |
| Urbanicity type |  |  |  |  |
| City | 12.1 | 1,376 | 12.1 | 1,376 |
| Combined Urban-Rural Areas | 3.4 | 387 | 3.4 | 387 |
| Town Center Areas | 13.2 | 1,500 | 13.2 | 1,500 |
| Combined Town-Township Areas | 8.9 | 1,008 | 8.9 | 1,008 |
| Special District | 0.7 | 79 | 0.7 | 79 |
| Township Center Areas | 3.9 | 442 | 3.9 | 442 |
| Village | 57.7 | 6,545 | 57.7 | 6,545 |
| Region |  |  |  |  |
| Beijing-Tianjin-Hebei-Shandong | 14.9 | 1,622 | 15.1 | 1,711 |
| Yangtze River Delta | 9.3 | 1,020 | 9.0 | 1,020 |
| SouthEast | 7.8 | 8,477 | 7.5 | 8,477 |
| Central | 29.9 | 3,264 | 28.8 | 3,264 |
| SouthWest | 21.2 | 2,320 | 23.4 | 2,649 |
| NorthWest | 9.2 | 1,002 | 8.8 | 1,002 |
| NorthEast | 7.7 | 844 | 7.4 | 844 |

Note: SD = standard deviation

### Life Satisfaction

LS was collected by asking “Please think about your life as a whole. How satisfied are you with it?” and the response could be completely satisfied (LS=5), very satisfied, somewhat satisfied, not very satisfied, or not at all satisfied (LS=1).

### Sleep

Respondents were asked about the hours of their actual sleep for one night (self-reported nighttime sleep duration) and duration of nap after lunch (self-reported afternoon napping time) during the past month, with truncation at 1% and 99% percentiles to eliminate outliers. Total sleep duration was obtained by summing up self-reported nighttime sleep duration and afternoon napping time. The subjects were asked to indicate the frequency for restlessness during sleep on a 4-item scale. If it happens most or all of the time, they got 4 for sleep quality (very poor). If it never happens, they got 1 for sleep quality (very good).

### Depression

Depression was measured by the 10-question version of the Center for Epidemiological Survey - Depression (CESD) (Zhao et al., 2020). The Cronbach’s alpha of the CESD-10 was 0.791. CESD scores range from 0 to 30, with higher score indicating more severe depressive symptoms.

### Sunset time

Sunset time and sunrise time were computed with the calculator provided by US National Oceanic and Atmospheric Administration. Annual average sunset time and month-specific sunset time for each city were recoded so that the integer digit represented the hour, and decimal digits the minutes divided by one hour. Hence, its coefficient represents the marginal effect of the effect of one additional hour.

### Covariates

Demographic covariates included age, gender, *hukou*, marital status (married or not), employment (non-agricultural employed, self-employed, agricultural, unemployed), years of schooling, households’ per capita expenditure (PCE), social participation (yes or no for social activities in the past month), and physical disability. PCE was imputed with the mean value of PCE by province, *hukou*, and gender. Physical disability was quantified by having difficulty with activities of daily living (ADL) or instrumental activities of daily living (IADL). Community level covariates were terrain and urbanicity type; city level covariates were GDP and population. Region by economic development level was also used as a covariate.

### Statistical Analysis

To obtain the effect of sleep duration on LS and mediation effect of depression in a stepwise way, we employed the Stata command feologit, a user-developed command for fixed-effects ordered logit model (Baetschmann et al., 2020). We computed direct and indirect effect of depression with bootstrap method on pooled data. Nonlinear relationships and heterogeneity were examined with feologit. For longitudinal analysis, time-invariant controls were dropped, such as gender and control variables at community and higher levels. All standard errors were clustered at the city level.

To further determine the source of endogeneity, we used instrumental variable ordered probit model (IV-oprobit) with the Stata command cmp (Roodman, 2011) and annual average sunset time as the instrument of sleep by *hukou* and employment (Gibson & Shrader, 2018; Giuntella et al., 2017; Giuntella & Mazzonna, 2019). Causal mediation analysis was conducted for employed population with urban *hukou*, using average sunset time for a pooled baseline model and month-specific sunset time for a fixed-effects model exploring seasonal variations.

Finally, we used information value (binary dependent variable) and random forest (ordered dependent variable) for predictive power of sleep measures on life satisfaction with R x64 4.0.3. Information value considers each variable’s independent contribution to the outcome and could be used to compare the strength of continuous and categorical variables without creating dummy variables (Kim, 2016). An information value great than 0.3 suggests strong predictive power by rule of thumb (Lin, 2013). Because categorical variables and continuous variables coexist, we computed importance ranking by mean decrease in accuracy criterion for random forest model (Zhang et al., 2018).

## Results

### Mediation, Non-linear relationship, and heterogeneity analysis

Table 3 shows effect of sleep duration on LS and the mediating role of depression with fixed-effects model. Effect of sleep hours on LS (OR=0.057, SE=0.018, p<0.01) is insignificant (OR=0.024, SE=0.018, p>0.05) after adding the mediator CESD (OR=-0.081, SE=0.005, p<0.001) negatively associated with sleep hours (B=-0.682, SE=0.030, p<0.001). Effect of total sleep duration on LS (OR=0.062, SE=0.015, p<0.001) decreases (OR=0.032, SE=0.015, p<0.05) after adding the mediator CESD (OR=-0.081, SE=0.005, p<0.001) negatively associated with total sleep duration (B=-0.597, SE=0.027, p<0.001). Effect of sleep quality on LS (OR=0.153, SE=0.027, p<0.001) is insignificant (OR=-0.049, SE=0.027, p>0.05) after adding the mediator CESD (OR=-0.091, SE=0.005, p<0.001) negatively associated with sleep quality (B=-2.478, SE=0.044, p<0.001).

**Table 3.** Sleep measures, depression, and life satisfaction (fixed-effects).

|  | LS | CESD | LS | LS | CESD | LS | LS | CESD | LS |
| --- | --- | --- | --- | --- | --- | --- | --- | --- | --- |
| Sleep Hours | 0.057**<br>(0.018) | -0.682***<br>(0.030) | 0.024<br>(0.018) |  |  |  |  |  |  |
| Total Sleep Duration |  |  |  | 0.062***<br>(0.015) | -0.597***<br>(0.027) | 0.032*<br>(0.015) |  |  |  |
| Sleep Quality |  |  |  |  |  |  | 0.153***<br>(0.027) | -2.478***<br>(0.044) | -0.049<br>(0.027) |
| CESD |  |  | -0.081***<br>(0.005) |  |  | -0.081***<br>(0.005) |  |  | -0.091***<br>(0.005) |
| N | 12,718 | 22,211 | 12,718 | 12,718 | 22,211 | 12,718 | 12,532 | 22,074 | 12,532 |
| Pseudo R Squared | 0.059 |  | 0.102 | 0.060 |  | 0.103 | 0.063 |  | 0.105 |

Note: Standard errors in parentheses, clustered at city level; odds ratio (OR) instead of coefficients; LS = life satisfaction; CESD = Center for Epidemiological Survey - Depression. Controls include *hukou*, marital status, years of schooling, expenditure per capita within households, age and age squared, physical disability, social participation, GDP and population at city level, and interview time. \* $p < 0.05$ , \*\* $p < 0.01$ , \*\*\* $p < 0.001$

Table 4 demonstrates mediation effect of depression in a pooled analysis. The indirect effect of sleep hours on LS is 0.032 (SE=0.001, p<0.001), while the direct effect is 0.012 (SE=0.003, p<0.001). For total sleep duration, indirect effect is 0.028 (SE=0.001, p<0.001) and the direct effect is 0.012 (SE=0.015, p<0.001). For sleep quality, indirect effect LS is 0.119 (SE=0.003, p<0.001) and direct effect is -0.018 (SE=0.005, p<0.001). Confidence interval of bias-corrected bootstrapping (replications set at 5,000) for the above effects does not include zero. Therefore, the conducive effect of sleep hours, total sleep duration, and sleep quality on LS mediated by depression is 72.7%, 65.1%, 117.8%, respectively.

**Table 4.** Mediating effect of depression (pooled)

|  | Point | Product of Coefficient |  | Bootstrapping |  |
| --- | --- | --- | --- | --- | --- |
|  | Estimate | SE | Z | Lower | Upper |
| *Sleep Hours* |  |  |  |  |  |
| Indirect | 0.032*** | 0.001 | 25.760 | 0.030 | 0.035 |
| Direct | 0.012*** | 0.003 | 4.004 | 0.006 | 0.018 |
| *Total Sleep Duration* |  |  |  |  |  |
| Indirect | 0.028*** | 0.001 | 24.987 | 0.025 | 0.030 |
| Direct | 0.015*** | 0.003 | 5.590 | 0.010 | 0.020 |
| *Sleep Quality* |  |  |  |  |  |
| Indirect | 0.119*** | 0.003 | 36.997 | 0.112 | 0.125 |
| Direct | -0.018*** | 0.005 | -3.744 | -0.028 | -0.009 |

Note: SE = standard error; bias-corrected (BC) bootstrap confidence interval; 5000 bootstrap replications, \*\*\* p<0.01

Supplementary Table 1 displays nonlinear relationship by adding squared terms of sleep hours and total sleep duration. No significant quadratic relationship for sleep hours (OR=-0.012, SE=0.006, p>0.05) or total sleep duration (OR=-0.003, SE=0.005, p>0.05) was found.

Supplementary Table 2 presents gender and age heterogeneity. While no gender-based difference was detected, effects of sleep hours (OR=0.093, SE=0.032, p<0.01), total sleep duration (OR=0.092, SE=0.030, p<0.01), and sleep quality (OR=0.205, SE=0.051, p<0.001) are the most significant among people aged 53-to-61 years. Supplementary Table 3 presents educational heterogeneity. Effects of sleep hours (OR=0.143, SE=0.046, p<0.01) and total sleep duration (OR=0.140, SE=0.039, p<0.001) are most significant among those only completed primary school, while effect of sleep quality (OR=0.192, SE=0.060, p<0.01) is the most significant among the illiterate population.

### Instrumental Variable Strategy and Source of Endogeneity

Table 5 and 6 present endogeneity among non-agricultural employed, and self-employed population respectively. Endogeneity is evident only among non-agricultural employed population with urban *hukou* (sleep hours, atanhrho12=-1,530, SE=0.757, p<0.05; total sleep duration, atanhrho=-1.848, SE=0.915, p<0.05) and sleep measures significantly impact LS (sleep hours, B=0.708, SE=0.071, p<0.001; total sleep duration, B=0.642, SE=0.040, p<0.001), but the effect of sunset is insignificant (p>0.5). Instead, only among self-employed population with urban *hukou*, coefficients of sunset (sleep hours, B=1.217, SE=0.290, p<0.001; total sleep duration, B=1.155, SE=0.296, p<0.001) passed the weak instrument test (sleep hours, Z=4.19, F=17.6>10; total sleep duration, Z=3.91, F=15.3>10). Among the agricultural and the unemployed population (Supplementary Table 4 and 5), although no endogeneity or relevant instrument was discovered, sleep duration significantly impacts rural population (farmer, B=0.443, SE=0.166, p<0.01; no work, B=-0.507, SE=0.093, p<0.001). Supplementary Figure 1-8 portray binned scatter plots of sleep duration and sunset after controlling for employment, *hukou*, and survey wave. By observation, the direction of correlation between sleep hours and sunset changed for non-agricultural employed population (by survey wave), self-employed and farmer population (by *hukou*). Supplementary Table 6 and 7 further investigate mediation effects with instrumental variable, but no significant effect was found (p>0.05).

**Table 5.** Endogeneity among Non-agricultural employed population (by *hukou*)

|  | Sleep Hours |  | Total Sleep Duration |  |
| --- | --- | --- | --- | --- |
|  | Rural | Urban | Rural | Urban |
| *LS* |  |  |  |  |
| Sleep Measures | 0.121<br>(0.103) | 0.708***<br>(0.071) | -0.405<br>(0.663) | 0.642***<br>(0.040) |
| *Sleep Measures* |  |  |  |  |
| sunset | -0.030<br>(0.152) | -0.239<br>(0.193) | 0.070<br>(0.192) | -0.196<br>(0.209) |
| constant | 14.737***<br>(3.372) | 13.139*<br>(5.390) | 12.888**<br>(4.217) | 13.438*<br>(6.150) |
| lnsigma | 0.435***<br>(0.013) | 0.314***<br>(0.031) | 0.559***<br>(0.014) | 0.438***<br>(0.029) |
| atanhrho12 | -0.108<br>(0.160) | -1.530*<br>(0.757) | 1.012<br>(2.539) | -1.848*<br>(0.915) |
| N | 4,275 | 1,185 | 4,275 | 1,185 |

Note: Standard errors in parentheses, clustered at city level; LS = life satisfaction. Controls include *hukou*, gender, marital status, years of schooling, expenditure per capita within households, age and age squared, physical disability, social participation, terrain and urbanicity type at community level, GDP and population at city level, region, and interview time. \* $p < 0.05$ , \*\* $p < 0.01$ , \*\*\* $p < 0.001$

**Table 6.**
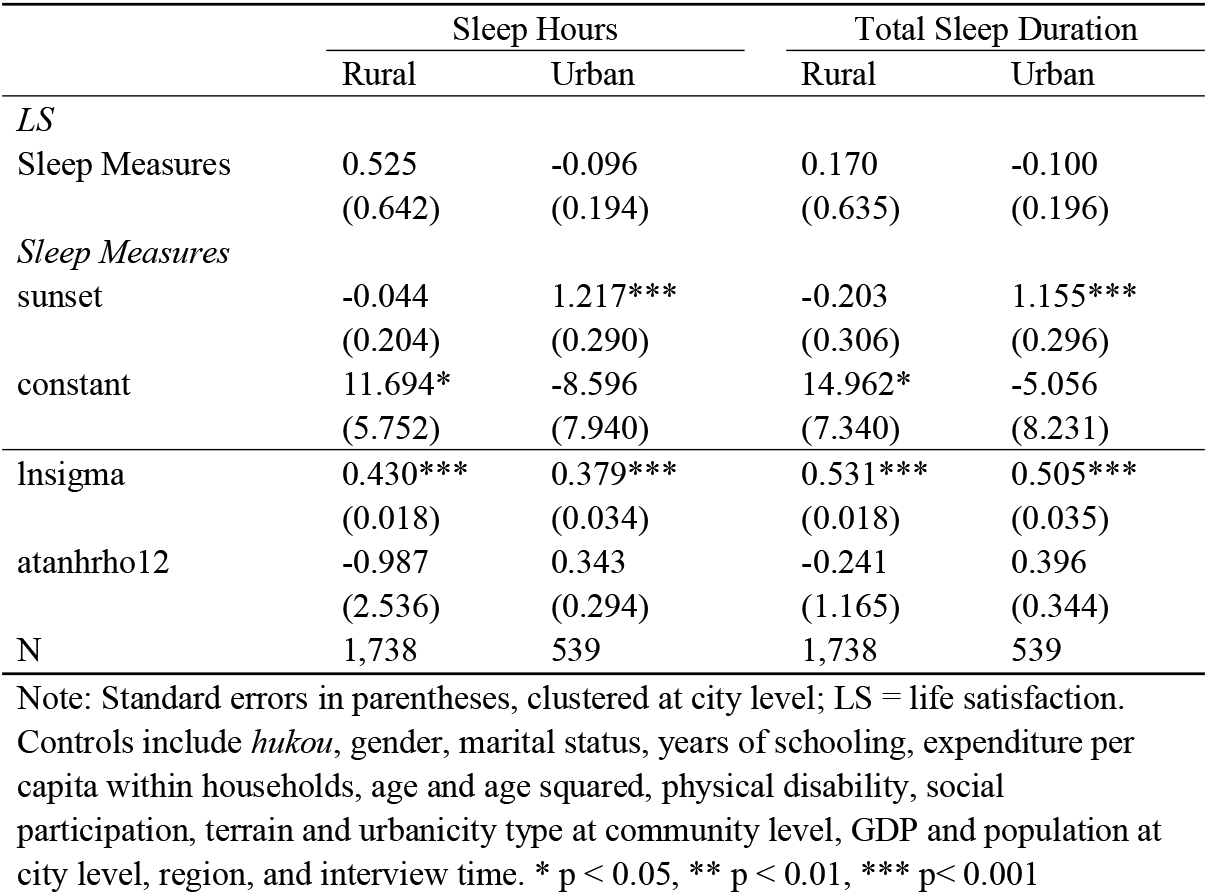
Endogeneity among self-employed population (by *hukou*)

|  | Sleep Hours |  | Total Sleep Duration |  |
| --- | --- | --- | --- | --- |
|  | Rural | Urban | Rural | Urban |
| *LS* |  |  |  |  |
| Sleep Measures | 0.525<br>(0.642) | -0.096<br>(0.194) | 0.170<br>(0.635) | -0.100<br>(0.196) |
| *Sleep Measures* |  |  |  |  |
| sunset | -0.044<br>(0.204) | 1.217***<br>(0.290) | -0.203<br>(0.306) | 1.155***<br>(0.296) |
| constant | 11.694*<br>(5.752) | -8.596<br>(7.940) | 14.962*<br>(7.340) | -5.056<br>(8.231) |
| lnsigma | 0.430***<br>(0.018) | 0.379***<br>(0.034) | 0.531***<br>(0.018) | 0.505***<br>(0.035) |
| atanhrho12 | -0.987<br>(2.536) | 0.343<br>(0.294) | -0.241<br>(1.165) | 0.396<br>(0.344) |
| N | 1,738 | 539 | 1,738 | 539 |

Note: Standard errors in parentheses, clustered at city level; LS = life satisfaction. Controls include *hukou*, gender, marital status, years of schooling, expenditure per capita within households, age and age squared, physical disability, social participation, terrain and urbanicity type at community level, GDP and population at city level, region, and interview time. \* $p < 0.05$ , \*\* $p < 0.01$ , \*\*\* $p < 0.001$

**Table 7.**
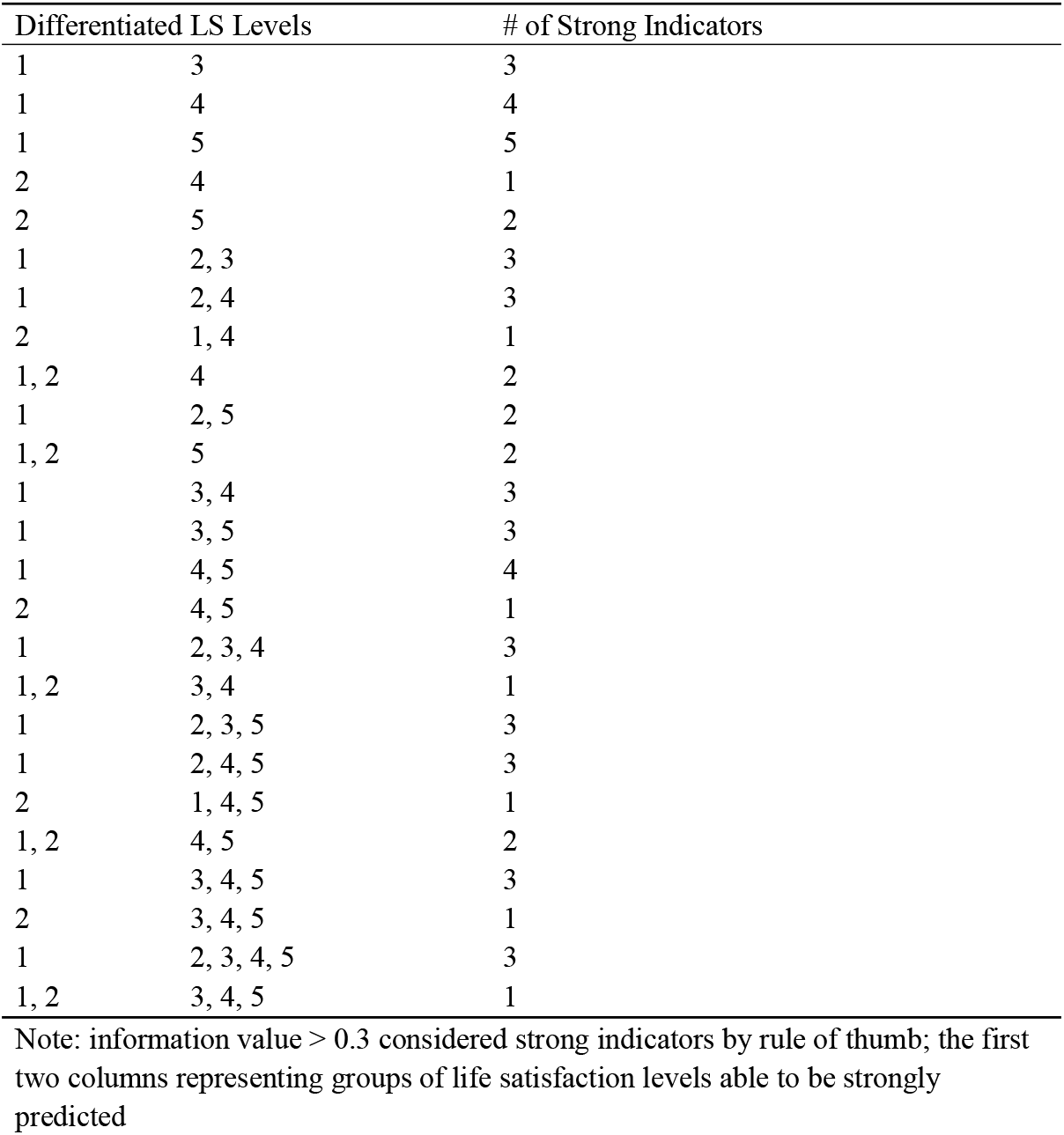
Predictive power of sleep measures as information value

### Predictive Power

Table 7 summarizes predictions by sleep measures. LS levels from the first column are strongly distinguished from those in the second column, and the third column gives the number of strong predictors. By observation, sleep measures differentiated LS level 1 and 2, representing poor life satisfaction, from good life satisfaction. Detailed information values on all LS combinations are presented in Supplementary Table 8. Supplementary Table 9 shows random forest ranking of predictive power for all variables. Depression, city-level GDP, and years of schooling ranked high, followed by city-level population, sleep quality, and nighttime sleep duration.

## Discussion

Using the 2015- and 2018-wave data of CHARLS, we found the protective effect of sleep duration and sleep quality on LS, largely mediated by depression. Specifically, good sleep promotes mental health, which in turn contributes to LS. The source of endogeneity with self-reported sleep measures is non-agricultural employed population with urban *hukou*. Poor LS can be predicted by self-reported sleep measures.

### Sleep, Life Satisfaction, and Depression as a Mediator

After controlling individual and time fixed effects, we found positive effects of sleep hours, total sleep duration, and sleep quality on LS. H1 was therefore supported. This is consistent with previous research which report that better sleep is associated with higher level of LS (Liu et al., 2020; Ness & Saksvik-Lehouillier, 2018; Piper, 2016) and may constitute part of LS intervention program (Diener & Biswas-Diener, 2019; Liu et al., 2020). Nonlinear relationship was found to be insignificant in the study, inconsistent with the U-shaped correlation reported in cross-sectional literature (Pagan, 2017; Yokoyama et al., 2008; Zhi et al., 2016). Respondents aged 53-to-61 years old with primary-school education mainly contributed to the effect of sleep duration, while those illiterate accounted for the effect of sleep quality most.

Depression was found to mediate the effects of sleep hours and sleep quality on LS, while partially mediating the effect of total sleep duration; depression was negatively associated with sleep measures and LS. H2 was therefore supported. This finding is consistent with the cross-sectional literature on the mediating role of depression (Zhi et al., 2016), but our longitudinal analysis demonstrates that effects of sleep hours and sleep quality are fully mediated by depression. Although one study confirmed sleep quality as outcome or mediator of the association between depression and LS (Brandolim Becker et al., 2018; Duong, 2021), the cross-sectional design could not determine the direction of association, which might also explain why sleep quality became negatively associated with LS in our pooled analysis of mediation effects. While the mediation effect may be explained by sleep-caused daytime tiredness and bidirectional relationship between poor sleep and lower LS (Koivumaa-Honkanen et al., 2004; Liu et al., 2020; Magee et al., 2011; Nes et al., 2013), depression displayed full mediation in terms of sleep hours and sleep quality because our longitudinal model eliminated individual and time fixed effects. As for total sleep duration, partial mediation was detected, possibly due to its complex correlation with other factors (Chen et al., 2018; Lin, 2018; Sun & Lin, 2016) and complicated reasons (Duggan et al., 2018) for napping, which goes beyond the focus of our study.

### Sunset and Source of Endogeneity

Endogeneity was found among non-agricultural employed population with urban *hukou*. H3 was therefore supported. This is consistent with the hypothesis in the literature (Gibson & Shrader, 2018; Giuntella et al., 2017; Giuntella & Mazzonna, 2019): Human bodies are inclined to sleep according to the environment because of the circadian rhythms, and later sunset time could lead to later darkness and thus postponed bedtime (Roenneberg et al., 2007; Roenneberg & Merrow, 2007; Wright Jr et al., 2013); Employed people living in urban areas are faced with rigid social schedules, thus those with later sunset time may not be able to compensate for later bedtime by waking up later in the mornings. This gap between solar cues and social schedules means that a later sunset can have important effects on sleep duration (Giuntella et al., 2017). Nevertheless, relevance of sunset time with sleep failed the weak instrument test, except for self-employed population with urban *hukou*. Supplementary figures supported the explanation that association of sunset time with sleep duration could be explained by covariates. After all, sunset time used in the literature barely passed weak instrument test (Giuntella et al., 2017). The positive correlation found among urban self-employed population might attribute to occupations responding to neither solar cues nor social schedule, meaning these observations of working age subjects can freely plan their daily schedule. Among farmer and those unemployed, sleep duration is significantly associated with rural *Hukou* despite no endogeneity or relevant instrument. One possible explanation is that they mainly respond to solar cues. Still, to our best knowledge, no study has covered associations between solar cues, social schedule, and sleep, expect for the outcomes of rigid social schedule against solar cues (Giuntella et al., 2017).

### Sleep Predicting Poor Life Satisfaction

Sleep duration/quality strongly predicted poor LS, ranking after CESD, city-level indicators, and education. H4 was therefore supported. Previous studies focused on traditional indicators of objective and perceived socioeconomic status, health, mental well-being, and other conventional demographics (Sancho et al., 2020; Zhang et al., 2018). Our findings revealed depression as one major predictor, but also highlighted the significance of broader indicators, such as city-level economic development. Moreover, sleep was found to be a useful predictor though neglected.

### Limitations and Conclusion

There are several limitations that need to be acknowledged. First, sunset time were collected at city level, as we could not access data at community level, which might explain the weak relevance of the instrument variable. Second, although we determine the source of endogeneity as non-agricultural employed population with urban *hukou*, it remains unclear why other population demonstrated significant correlations with sunset under some circumstances. Relations between solar cues, social schedule, and sleep are needs further examination. Third, a considerable number of respondents reported both agricultural and non-agricultural occupations (4,383 of the total 5,780 non-agricultural employed participants also reported occupations as farmers), while we focused on main occupation. Future studies may consider a more specific classification of employment and the interaction between main occupation and other occupations.

Reliable findings on the relationship between sleep and LS require handling of the prevalent self-reported sleep measures in social surveys. In the current study, we confirmed the effect of sleep on life satisfaction with fixed-effects model, which is mediated by depression, and urban non-agricultural employed population as the source of endogeneity by instrumental variables. Researchers need to caution the differences between objectively measured and self-reported sleep measures. Although the predictive power of sleep measures did not rank high, they are useful in predicting poor life satisfaction. Policy makers may consider campaigns to promote healthy sleep habits and protection of mental health, while detecting the vulnerable population by sleep screening programs.

## Data Availability

All data produced are available online at official website of the China Health and Retirement Longitudinal Study (CHARLS): http://charls.pku.edu.cn/.

## Acknowledgments

We thank the China Health and Retirement Longitudinal Study (CHARLS) for collecting data and Peking University School of Health Humanities for financing this study.

## Availability of data and material

Data and related materials are available at official website of the China Health and Retirement Longitudinal Study (CHARLS): http://charls.pku.edu.cn/.

## Code availability

Stata/SE 16.0 and R x64 4.0.3 were employed for data analysis.

## Ethics approval and consent to participate

This study used the data from the China Health and Retirement Longitudinal Study (CHARLS), which has obtained the ethics approval and subjects’ consent to participate. Ethical approval for all CHARLS waves was granted from the Institutional Review Board at Peking University. The IRB approval number for the main household survey, including anthropometrics, is IRB00001052-11015; the IRB approval number for biomarker collection, was IRB00001052-11014. We consulted the ethics committee office at Peking University and was informed that no ethical review was required since we used a public database for extracting our data and no personal information was revealed in the article. Therefore, we ask for waiving of the ethics approval and consent to participate.

