## Supplementary Table 1-9, Supplementary Figure 1-8 for "Sun, Sleep, and Satisfaction: Mediating Role of Depression and Source of Endogeneity among Middle-aged and Older Adults in China"

| **Supplementary Table 1.** Nonlinear relationship between sleep duration and life satisfaction | | |
| --- | --- | --- |
|  | LS | LS |
| Sleep Hours | 0.206* |  |
|  | (0.082) |  |
| Sleep Hours Squared | -0.012 |  |
|  | (0.006) |  |
| Total Sleep Duration |  | 0.099 |
|  |  | (0.069) |
| Total Sleep Duration Squared |  | -0.003 |
|  |  | (0.005) |
| N | 12,718 | 12,718 |
| Pseudo R Squared | 0.059 | 0.060 |
| Note: Standard errors in parentheses, clustered at city level; odds ratio (OR) instead of coefficients; LS = life satisfaction. Controls include *hukou*, marital status, years of schooling, expenditure per capita within households, age and age squared, physical disability, social participation, GDP and population at city level, and interview time. * p < 0.05, ** p < 0.01, *** p< 0.001 | | |

| **Supplementary Table 2.** Heterogeneity with fixed-effects (gender & age) | | | | | | |
| --- | --- | --- | --- | --- | --- | --- |
|  | Gender | |  | Age | | |
|  | Female | Male |  | Aged 45-52 | Aged 53-61 | Aged 62-70 |
| *Sleep Hours* | 0.043* | 0.073** |  | 0.104* | 0.093** | 0.045 |
|  | (0.021) | (0.026) |  | (0.045) | (0.032) | (0.031) |
| N | 6,776 | 5,888 |  | 2,458 | 2,998 | 3,098 |
| Pseudo R Squared | 0.056 | 0.070 |  | 0.089 | 0.057 | 0.040 |
| *Total Sleep Duration* | 0.052** | 0.072*** |  | 0.089* | 0.092** | 0.062* |
|  | (0.019) | (0.021) |  | (0.038) | (0.030) | (0.026) |
| N | 6,776 | 5,888 |  | 2,458 | 2,998 | 3,098 |
| Pseudo R Squared | 0.057 | 0.071 |  | 0.088 | 0.058 | 0.042 |
| *Sleep Quality* | 0.136*** | 0.169*** |  | 0.034 | 0.205*** | 0.134** |
|  | (0.034) | (0.038) |  | (0.053) | (0.051) | (0.050) |
| N | 6,658 | 5,820 |  | 2,436 | 2,958 | 3,022 |
| Pseudo R Squared | 0.059 | 0.075 |  | 0.085 | 0.062 | 0.042 |
| Note: Standard errors in parentheses, clustered at city level; odds ratio (OR) instead of coefficients; LS = life satisfaction; CESD = Center for Epidemiological Survey - Depression. Controls include *hukou*, marital status, years of schooling, expenditure per capita within households, age and age squared, physical disability, social participation, GDP and population at city level, and interview time. * p < 0.05, ** p < 0.01, *** p< 0.001 | | | | | | |

| **Supplementary Table 3.** Heterogeneity with fixed-effects (education) | | | | | |
| --- | --- | --- | --- | --- | --- |
|  | Illiterate | Did not finish primary | Finished primary | Middle school | High school and above |
| *Sleep Hours* | -0.032 | 0.076 | 0.143** | 0.007 | 0.144 |
|  | (0.032) | (0.045) | (0.046) | (0.050) | (0.074) |
| N | 2,290 | 1,946 | 1,986 | 2,498 | 1,364 |
| Pseudo R Squared | 0.070 | 0.091 | 0.079 | 0.073 | 0.079 |
| *Total Sleep Duration* | -0.009 | 0.085* | 0.140*** | 0.017 | 0.121 |
|  | (0.031) | (0.038) | (0.039) | (0.044) | (0.068) |
| N | 2,290 | 1,946 | 1,986 | 2,498 | 1,364 |
| Pseudo R Squared | 0.069 | 0.093 | 0.081 | 0.073 | 0.078 |
| *Sleep Quality* | 0.192** | 0.114* | 0.153* | 0.131* | 0.161* |
|  | (0.060) | (0.058) | (0.068) | (0.062) | (0.081) |
| N | 2,254 | 1,916 | 1,952 | 2,476 | 1,364 |
| Pseudo R Squared | 0.079 | 0.091 | 0.076 | 0.075 | 0.078 |
| Note: Standard errors in parentheses, clustered at city level; odds ratio (OR) instead of coefficients; LS = life satisfaction; CESD = Center for Epidemiological Survey - Depression. Controls include *hukou*, marital status, years of schooling, expenditure per capita within households, age and age squared, physical disability, social participation, GDP and population at city level, and interview time. * p < 0.05, ** p < 0.01, *** p< 0.001 | | | | | |

| **Supplementary Table 4.** Endogeneity among farmer population (by *hukou*) | | | | | |
| --- | --- | --- | --- | --- | --- |
|  | Sleep Hours | |  | Total Sleep Duration | |
|  | Rural | Urban |  | Rural | Urban |
| *LS* |  |  |  |  |  |
| Sleep Measures | 0.443** | 0.591 |  | 0.262 | -0.493 |
|  | (0.166) | (0.399) |  | (0.166) | (0.289) |
| *Sleep Measures* |  |  |  |  |  |
| sunset | -0.134 | 0.206 |  | -0.302* | -0.181 |
|  | (0.101) | (0.411) |  | (0.147) | (0.389) |
| constant | 14.276*** | 2.117 |  | 18.841*** | 12.528 |
|  | (2.538) | (10.657) |  | (3.504) | (11.341) |
| lnsigma | 0.599*** | 0.394*** |  | 0.708*** | 0.532*** |
|  | (0.012) | (0.033) |  | (0.010) | (0.032) |
| atanhrho12 | -0.932 | -1.082 |  | -0.453 | 1.432 |
|  | (0.730) | (1.962) |  | (0.432) | (1.954) |
| N | 8,053 | 484 |  | 8,053 | 484 |
| Note: Standard errors in parentheses, clustered at city level; LS = life satisfaction. Controls include *hukou*, gender, marital status, years of schooling, expenditure per capita within households, age and age squared, physical disability, social participation, Terrain and Urbanicity at community level, GDP and population at city level, region, and interview time. * p < 0.05, ** p < 0.01, *** p< 0.001 | | | | | |

| **Supplementary Table 5.** Endogeneity among no work population (by *hukou*) | | | | | |
| --- | --- | --- | --- | --- | --- |
|  | Sleep Hours | |  | Total Sleep Duration | |
|  | Rural | Urban |  | Rural | Urban |
| *LS* |  |  |  |  |  |
| Sleep Measures | -0.507*** | 0.395 |  | 0.047 | 0.424 |
|  | (0.093) | (0.292) |  | (0.734) | (0.236) |
| *Sleep Measures* |  |  |  |  |  |
| sunset | 0.096 | 0.440* |  | 0.030 | 0.372 |
|  | (0.255) | (0.190) |  | (0.878) | (0.208) |
| constant | 6.141 | 12.486* |  | 10.446 | 17.380** |
|  | (5.806) | (5.055) |  | (17.893) | (5.630) |
| lnsigma | 0.635*** | 0.418*** |  | 0.750*** | 0.547*** |
|  | (0.015) | (0.020) |  | (0.015) | (0.021) |
| atanhrho12 | 2.322 | -0.538 |  | 0.040 | -0.717 |
|  | (2.834) | (0.644) |  | (1.553) | (0.755) |
| N | 3,114 | 2,067 |  | 3,114 | 2,067 |
| Note: Standard errors in parentheses, clustered at city level; LS = life satisfaction. Controls include hukou, gender, marital status, years of schooling, expenditure per capita within households, age and age squared, physical disability, social participation, Terrain and Urbanicity at community level, GDP and population at city level, region, and interview time. * p < 0.05, ** p < 0.01, *** p< 0.001 | | | | | |

| **Supplementary Table 6.** Mediation effect of depression using instrumental variable strategy (urban non-agricultural employed population) | | | | | |
| --- | --- | --- | --- | --- | --- |
|  | Sleep Hours | |  | Total Sleep Duration | |
|  | Baseline | Seasonal |  | Baseline | Seasonal |
| Total Effect | 1.024 | 0.091 |  | 1.248 | 0.110 |
|  | (0.801) | (0.314) |  | (1.162) | (0.367) |
| Direct Effect | -0.123 | 0.088 |  | -0.086 | 0.102 |
|  | (0.129) | (0.324) |  | (0.092) | (0.423) |
| Indirect Effect | 1.147 | 0.004 |  | 1.334 | 0.008 |
|  | (1.295) | (0.106) |  | (1.599) | (0.245) |
| N | 1,185 | 1,214 |  | 1,185 | 1,214 |
| F (first stage one) | 1.488 | 2.133 |  | 0.853 | 1.300 |
| F (first stage two) | 1.369 | 0.099 |  | 1.585 | 0.045 |
| Individual Fixed Effect | No | Yes |  | No | Yes |
| Note: Standard errors in parentheses, clustered at city level; LS = life satisfaction. Controls include *hukou*, gender, marital status, years of schooling, expenditure per capita within households, age and age squared, physical disability, social participation, Terrain and Urbanicity at community level, GDP and population at city level, region, and interview time. * p < 0.05, ** p < 0.01, *** p< 0.001 | | | | | |

| **Supplementary Table 7.** Mediation effect of depression using instrumental variable strategy (urban self-employed population) | | | | | |
| --- | --- | --- | --- | --- | --- |
|  | Sleep Hours | |  | Total Sleep Duration | |
|  | Baseline | Seasonal |  | Baseline | Seasonal |
| Total Effect | -0.062 | -0.735 |  | -0.065 | -0.508 |
|  | (0.132) | (0.898) |  | (0.137) | (0.466) |
| Direct Effect | -0.010 | -0.126 |  | -0.008 | -0.101 |
|  | (0.106) | (0.213) |  | (0.109) | (0.178) |
| Indirect Effect | -0.052 | -0.609 |  | -0.057 | -0.407 |
|  | (0.135) | (1.036) |  | (0.152) | (0.604) |
| N | 539 | 550 |  | 539 | 550 |
| F (first stage one) | 16.514 | 0.505 |  | 14.375 | 1.217 |
| F (first stage two) | 1.255 | 0.704 |  | 1.091 | 0.852 |
| Individual Fixed Effect | No | Yes |  | No | Yes |
| Note: Standard errors in parentheses, clustered at city level; LS = life satisfaction. Controls include *hukou*, gender, marital status, years of schooling, expenditure per capita within households, age and age squared, physical disability, social participation, Terrain and Urbanicity at community level, GDP and population at city level, region, and interview time. * p < 0.05, ** p < 0.01, *** p< 0.001 | | | | | |

| **Supplementary Table 8.** Information values of sleep measures on binary indicators of life satisfaction | | | | | | | | | | | | | | |
| --- | --- | --- | --- | --- | --- | --- | --- | --- | --- | --- | --- | --- | --- | --- |
| Life Satisfaction | Sleep Quality | |  | Sleep Hours | |  | Sleep Hours ≥ 7 | |  | Napping Time | |  | Nap or not | |
|  | 2015 | 2018 |  | 2015 | 2018 |  | 2015 | 2018 |  | 2015 | 2018 |  | 2015 | 2018 |
| 1 or 2 | 0.113 | 0.104 |  | 0.184 | 0.109 |  | 0.065 | 0.003 |  | 0.003 | 0.011 |  | 0.001 | 0.003 |
| 1 or 3 | 0.425 | 0.499 |  | 0.376 | 0.253 |  | 0.168 | 0.062 |  | 0.049 | 0.057 |  | 0.030 | 0.045 |
| 1 or 4 | 0.651 | 0.794 |  | 0.487 | 0.390 |  | 0.270 | 0.155 |  | 0.064 | 0.068 |  | 0.050 | 0.061 |
| 1 or 5 | 0.923 | 0.823 |  | 0.646 | 0.361 |  | 0.352 | 0.139 |  | 0.058 | 0.087 |  | 0.064 | 0.080 |
| 2 or 3 | 0.129 | 0.223 |  | 0.054 | 0.085 |  | 0.024 | 0.036 |  | 0.031 | 0.031 |  | 0.020 | 0.024 |
| 2 or 4 | 0.270 | 0.496 |  | 0.110 | 0.191 |  | 0.070 | 0.112 |  | 0.055 | 0.072 |  | 0.037 | 0.036 |
| 2 or 5 | 0.436 | 0.519 |  | 0.171 | 0.185 |  | 0.114 | 0.098 |  | 0.050 | 0.043 |  | 0.049 | 0.051 |
| 3 or 4 | 0.029 | 0.062 |  | 0.017 | 0.028 |  | 0.012 | 0.021 |  | 0.012 | 0.013 |  | 0.002 | 0.001 |
| 3 or 5 | 0.096 | 0.071 |  | 0.042 | 0.019 |  | 0.034 | 0.015 |  | 0.007 | 0.007 |  | 0.006 | 0.005 |
| 4 or 5 | 0.026 | 0.003 |  | 0.008 | 0.012 |  | 0.005 | 0.000 |  | 0.014 | 0.020 |  | 0.001 | 0.001 |
| 1 or 2 3 | 0.375 | 0.422 |  | 0.346 | 0.221 |  | 0.154 | 0.051 |  | 0.048 | 0.043 |  | 0.025 | 0.037 |
| 2 or 1 3 | 0.116 | 0.193 |  | 0.044 | 0.073 |  | 0.021 | 0.032 |  | 0.029 | 0.026 |  | 0.019 | 0.021 |
| 3 or 1 2 | 0.165 | 0.275 |  | 0.086 | 0.117 |  | 0.040 | 0.042 |  | 0.034 | 0.028 |  | 0.022 | 0.029 |
| 1 or 2 4 | 0.526 | 0.547 |  | 0.421 | 0.260 |  | 0.229 | 0.102 |  | 0.047 | 0.044 |  | 0.037 | 0.041 |
| 2 or 1 4 | 0.237 | 0.392 |  | 0.093 | 0.145 |  | 0.060 | 0.090 |  | 0.051 | 0.061 |  | 0.033 | 0.028 |
| 4 or 1 2 | 0.322 | 0.555 |  | 0.155 | 0.224 |  | 0.096 | 0.122 |  | 0.056 | 0.062 |  | 0.039 | 0.042 |
| 1 or 2 5 | 0.386 | 0.250 |  | 0.331 | 0.140 |  | 0.179 | 0.032 |  | 0.019 | 0.042 |  | 0.020 | 0.020 |
| 2 or 1 5 | 0.219 | 0.187 |  | 0.076 | 0.054 |  | 0.053 | 0.033 |  | 0.032 | 0.022 |  | 0.030 | 0.015 |
| 5 or 1 2 | 0.508 | 0.580 |  | 0.224 | 0.205 |  | 0.147 | 0.108 |  | 0.051 | 0.047 |  | 0.052 | 0.058 |
| 1 or 3 4 | 0.509 | 0.583 |  | 0.417 | 0.284 |  | 0.207 | 0.088 |  | 0.041 | 0.058 |  | 0.038 | 0.050 |
| 3 or 1 4 | 0.020 | 0.038 |  | 0.012 | 0.018 |  | 0.008 | 0.012 |  | 0.011 | 0.011 |  | 0.002 | 0.000 |
| 4 or 1 3 | 0.035 | 0.074 |  | 0.020 | 0.033 |  | 0.015 | 0.024 |  | 0.013 | 0.013 |  | 0.003 | 0.002 |
| 1 or 3 5 | 0.468 | 0.519 |  | 0.395 | 0.259 |  | 0.185 | 0.067 |  | 0.033 | 0.059 |  | 0.033 | 0.048 |
| 3 or 1 5 | 0.014 | 0.049 |  | 0.016 | 0.032 |  | 0.006 | 0.000 |  | 0.003 | 0.004 |  | 0.001 | 0.001 |
| 5 or 1 3 | 0.108 | 0.084 |  | 0.048 | 0.024 |  | 0.038 | 0.018 |  | 0.008 | 0.009 |  | 0.007 | 0.007 |
| 1 or 4 5 | 0.688 | 0.798 |  | 0.502 | 0.383 |  | 0.282 | 0.152 |  | 0.062 | 0.069 |  | 0.052 | 0.064 |
| 4 or 1 5 | 0.003 | 0.107 |  | 0.011 | 0.075 |  | 0.001 | 0.023 |  | 0.007 | 0.016 |  | 0.000 | 0.005 |
| 5 or 1 4 | 0.038 | 0.013 |  | 0.013 | 0.011 |  | 0.009 | 0.000 |  | 0.005 | 0.020 |  | 0.002 | 0.003 |
| 2 or 3 4 | 0.179 | 0.299 |  | 0.072 | 0.114 |  | 0.040 | 0.057 |  | 0.036 | 0.042 |  | 0.026 | 0.028 |
| 3 or 2 4 | 0.009 | 0.013 |  | 0.007 | 0.008 |  | 0.005 | 0.005 |  | 0.008 | 0.005 |  | 0.000 | 0.000 |
| 4 or 2 3 | 0.044 | 0.091 |  | 0.022 | 0.040 |  | 0.016 | 0.028 |  | 0.014 | 0.017 |  | 0.004 | 0.003 |
| 2 or 3 5 | 0.152 | 0.241 |  | 0.063 | 0.090 |  | 0.031 | 0.040 |  | 0.033 | 0.033 |  | 0.023 | 0.026 |
| 3 or 2 5 | 0.006 | 0.043 |  | 0.008 | 0.023 |  | 0.000 | 0.005 |  | 0.006 | 0.009 |  | 0.001 | 0.005 |
| 5 or 2 3 | 0.122 | 0.102 |  | 0.053 | 0.029 |  | 0.040 | 0.022 |  | 0.013 | 0.010 |  | 0.009 | 0.008 |
| 2 or 4 5 | 0.291 | 0.499 |  | 0.118 | 0.188 |  | 0.076 | 0.110 |  | 0.054 | 0.067 |  | 0.038 | 0.038 |
| 4 or 2 5 | 0.045 | 0.182 |  | 0.018 | 0.085 |  | 0.009 | 0.046 |  | 0.018 | 0.038 |  | 0.007 | 0.011 |
| 5 or 2 4 | 0.056 | 0.032 |  | 0.019 | 0.010 |  | 0.013 | 0.003 |  | 0.005 | 0.015 |  | 0.004 | 0.006 |
| 3 or 4 5 | 0.036 | 0.063 |  | 0.020 | 0.026 |  | 0.015 | 0.020 |  | 0.011 | 0.010 |  | 0.003 | 0.002 |
| 4 or 3 5 | 0.020 | 0.052 |  | 0.011 | 0.025 |  | 0.008 | 0.018 |  | 0.011 | 0.013 |  | 0.002 | 0.001 |
| 5 or 3 4 | 0.060 | 0.034 |  | 0.038 | 0.009 |  | 0.019 | 0.006 |  | 0.003 | 0.008 |  | 0.004 | 0.004 |
| 1 or 2 3 4 | 0.468 | 0.515 |  | 0.394 | 0.254 |  | 0.194 | 0.077 |  | 0.036 | 0.052 |  | 0.033 | 0.044 |
| 2 or 1 3 4 | 0.169 | 0.274 |  | 0.066 | 0.102 |  | 0.037 | 0.052 |  | 0.035 | 0.040 |  | 0.025 | 0.026 |
| 3 or 1 2 4 | 0.006 | 0.013 |  | 0.006 | 0.009 |  | 0.003 | 0.002 |  | 0.007 | 0.006 |  | 0.000 | 0.001 |
| 4 or 1 2 3 | 0.050 | 0.104 |  | 0.025 | 0.045 |  | 0.019 | 0.031 |  | 0.015 | 0.017 |  | 0.005 | 0.004 |
| 1 2 or 3 4 | 0.222 | 0.354 |  | 0.110 | 0.144 |  | 0.060 | 0.064 |  | 0.037 | 0.040 |  | 0.028 | 0.033 |
| 1 3 or 2 4 | 0.012 | 0.016 |  | 0.009 | 0.009 |  | 0.006 | 0.007 |  | 0.008 | 0.005 |  | 0.001 | 0.000 |
| 1 4 or 2 3 | 0.032 | 0.057 |  | 0.016 | 0.025 |  | 0.012 | 0.018 |  | 0.012 | 0.014 |  | 0.003 | 0.001 |
| 1 or 2 3 5 | 0.416 | 0.443 |  | 0.366 | 0.225 |  | 0.170 | 0.056 |  | 0.052 | 0.046 |  | 0.028 | 0.040 |
| 2 or 1 3 5 | 0.139 | 0.212 |  | 0.055 | 0.077 |  | 0.027 | 0.036 |  | 0.031 | 0.027 |  | 0.021 | 0.023 |
| 3 or 1 2 5 | 0.013 | 0.088 |  | 0.016 | 0.046 |  | 0.001 | 0.010 |  | 0.007 | 0.011 |  | 0.002 | 0.009 |
| 5 or 1 2 3 | 0.133 | 0.115 |  | 0.059 | 0.034 |  | 0.044 | 0.024 |  | 0.014 | 0.007 |  | 0.010 | 0.010 |
| 1 2 or 3 5 | 0.192 | 0.294 |  | 0.098 | 0.120 |  | 0.049 | 0.046 |  | 0.036 | 0.029 |  | 0.024 | 0.031 |
| 1 3 or 2 5 | 0.005 | 0.029 |  | 0.007 | 0.016 |  | 0.001 | 0.004 |  | 0.005 | 0.008 |  | 0.001 | 0.004 |
| 1 5 or 2 3 | 0.025 | 0.041 |  | 0.019 | 0.024 |  | 0.009 | 0.000 |  | 0.003 | 0.004 |  | 0.002 | 0.000 |
| 1 or 2 4 5 | 0.569 | 0.575 |  | 0.440 | 0.269 |  | 0.244 | 0.106 |  | 0.048 | 0.051 |  | 0.040 | 0.045 |
| 2 or 1 4 5 | 0.260 | 0.407 |  | 0.102 | 0.148 |  | 0.067 | 0.091 |  | 0.050 | 0.059 |  | 0.035 | 0.031 |
| 4 or 1 2 5 | 0.072 | 0.253 |  | 0.038 | 0.118 |  | 0.019 | 0.060 |  | 0.021 | 0.037 |  | 0.009 | 0.017 |
| 5 or 1 2 4 | 0.069 | 0.053 |  | 0.025 | 0.016 |  | 0.017 | 0.006 |  | 0.005 | 0.018 |  | 0.004 | 0.009 |
| 1 2 or 4 5 | 0.346 | 0.558 |  | 0.164 | 0.220 |  | 0.103 | 0.120 |  | 0.055 | 0.061 |  | 0.041 | 0.044 |
| 1 4 or 2 5 | 0.033 | 0.122 |  | 0.011 | 0.056 |  | 0.006 | 0.032 |  | 0.015 | 0.032 |  | 0.005 | 0.007 |
| 1 5 or 2 4 | 0.002 | 0.036 |  | 0.006 | 0.028 |  | 0.000 | 0.006 |  | 0.004 | 0.006 |  | 0.000 | 0.001 |
| 1 or 3 4 5 | 0.533 | 0.594 |  | 0.428 | 0.287 |  | 0.216 | 0.091 |  | 0.042 | 0.059 |  | 0.039 | 0.052 |
| 3 or 1 4 5 | 0.026 | 0.041 |  | 0.015 | 0.018 |  | 0.011 | 0.012 |  | 0.009 | 0.008 |  | 0.002 | 0.000 |
| 4 or 1 3 5 | 0.024 | 0.063 |  | 0.014 | 0.030 |  | 0.010 | 0.021 |  | 0.011 | 0.013 |  | 0.002 | 0.001 |
| 5 or 1 3 4 | 0.067 | 0.041 |  | 0.041 | 0.010 |  | 0.021 | 0.007 |  | 0.003 | 0.009 |  | 0.004 | 0.004 |
| 1 3 or 4 5 | 0.043 | 0.075 |  | 0.024 | 0.032 |  | 0.017 | 0.023 |  | 0.011 | 0.010 |  | 0.004 | 0.002 |
| 1 4 or 3 5 | 0.013 | 0.031 |  | 0.008 | 0.016 |  | 0.005 | 0.010 |  | 0.009 | 0.011 |  | 0.001 | 0.000 |
| 1 5 or 3 4 | 0.003 | 0.055 |  | 0.019 | 0.037 |  | 0.001 | 0.003 |  | 0.002 | 0.005 |  | 0.000 | 0.002 |
| 2 or 3 4 5 | 0.192 | 0.310 |  | 0.078 | 0.116 |  | 0.044 | 0.059 |  | 0.037 | 0.043 |  | 0.028 | 0.029 |
| 3 or 2 4 5 | 0.014 | 0.017 |  | 0.011 | 0.008 |  | 0.007 | 0.006 |  | 0.007 | 0.004 |  | 0.001 | 0.000 |
| 4 or 2 3 5 | 0.032 | 0.078 |  | 0.016 | 0.035 |  | 0.012 | 0.025 |  | 0.012 | 0.016 |  | 0.003 | 0.002 |
| 5 or 2 3 4 | 0.076 | 0.052 |  | 0.044 | 0.013 |  | 0.023 | 0.009 |  | 0.004 | 0.009 |  | 0.005 | 0.006 |
| 2 3 or 4 5 | 0.052 | 0.092 |  | 0.026 | 0.038 |  | 0.019 | 0.027 |  | 0.013 | 0.013 |  | 0.005 | 0.004 |
| 2 4 or 3 5 | 0.005 | 0.010 |  | 0.005 | 0.007 |  | 0.002 | 0.004 |  | 0.006 | 0.005 |  | 0.000 | 0.000 |
| 2 5 or 3 4 | 0.015 | 0.075 |  | 0.010 | 0.037 |  | 0.001 | 0.014 |  | 0.008 | 0.016 |  | 0.003 | 0.007 |
| 1 or 2 3 4 5 | 0.491 | 0.527 |  | 0.405 | 0.258 |  | 0.203 | 0.079 |  | 0.037 | 0.054 |  | 0.035 | 0.045 |
| 2 or 1 3 4 5 | 0.182 | 0.285 |  | 0.072 | 0.105 |  | 0.041 | 0.055 |  | 0.036 | 0.041 |  | 0.027 | 0.027 |
| 3 or 1 2 4 5 | 0.010 | 0.015 |  | 0.008 | 0.008 |  | 0.005 | 0.003 |  | 0.006 | 0.004 |  | 0.000 | 0.000 |
| 4 or 1 2 3 5 | 0.036 | 0.090 |  | 0.019 | 0.041 |  | 0.014 | 0.027 |  | 0.013 | 0.016 |  | 0.004 | 0.003 |
| 5 or 1 2 3 4 | 0.082 | 0.060 |  | 0.034 | 0.016 |  | 0.025 | 0.011 |  | 0.004 | 0.010 |  | 0.005 | 0.006 |
| 1 2 or 3 4 5 | 0.237 | 0.365 |  | 0.116 | 0.147 |  | 0.065 | 0.066 |  | 0.038 | 0.041 |  | 0.030 | 0.034 |
| 1 3 or 2 4 5 | 0.018 | 0.021 |  | 0.012 | 0.010 |  | 0.009 | 0.008 |  | 0.007 | 0.003 |  | 0.001 | 0.000 |
| 1 4 or 2 3 5 | 0.022 | 0.047 |  | 0.011 | 0.022 |  | 0.008 | 0.015 |  | 0.011 | 0.014 |  | 0.002 | 0.001 |
| 1 5 or 2 3 4 | 0.006 | 0.041 |  | 0.011 | 0.028 |  | 0.002 | 0.001 |  | 0.001 | 0.004 |  | 0.001 | 0.001 |
| 2 3 or 1 4 5 | 0.040 | 0.062 |  | 0.019 | 0.025 |  | 0.015 | 0.018 |  | 0.011 | 0.011 |  | 0.004 | 0.002 |
| 2 4 or 1 3 5 | 0.007 | 0.011 |  | 0.005 | 0.007 |  | 0.003 | 0.005 |  | 0.007 | 0.005 |  | 0.000 | 0.000 |
| 2 5 or 1 3 4 | 0.012 | 0.062 |  | 0.005 | 0.030 |  | 0.001 | 0.012 |  | 0.007 | 0.015 |  | 0.003 | 0.006 |
| 3 4 or 1 2 5 | 0.030 | 0.129 |  | 0.020 | 0.063 |  | 0.005 | 0.022 |  | 0.009 | 0.018 |  | 0.005 | 0.012 |
| 3 5 or 1 2 4 | 0.004 | 0.011 |  | 0.004 | 0.008 |  | 0.001 | 0.001 |  | 0.006 | 0.006 |  | 0.000 | 0.001 |
| 4 5 or 1 2 3 | 0.059 | 0.105 |  | 0.029 | 0.043 |  | 0.022 | 0.030 |  | 0.013 | 0.014 |  | 0.006 | 0.005 |

| **Supplementary Table 9.** Ranking of predictive power using random forest | | | |
| --- | --- | --- | --- |
| Variables | Panel | 2015 | 2018 |
| Sleep Quality | 5 | 5 | 6 |
| Sleep Hours | 6 | 6 | 7 |
| Napping Time | 14 | 11 | 24 |
| Sleep More than 7 Hours | 21 | 17 | 19 |
| Nap or not | 20 | 12 | 27 |
| Age | 12 | 7 | 12 |
| Urbanicity 112 | 34 | 35 | 34 |
| Urbanicity 121 | 31 | 26 | 29 |
| Urbanicity 122 | 26 | 33 | 31 |
| Urbanicity 123 | 35 | 34 | 35 |
| Urbanicity 210 | 32 | 25 | 33 |
| Urbanicity 220 | 9 | 9 | 8 |
| CESD | 1 | 1 | 1 |
| Disability | 16 | 18 | 9 |
| *Employment* |  |  |  |
| Self-employed | 33 | 28 | 32 |
| Farmer | 11 | 14 | 15 |
| No Work | 17 | 23 | 11 |
| Female | 19 | 20 | 26 |
| *Terrain* |  |  |  |
| Hill | 15 | 21 | 17 |
| Mountainous Region | 10 | 10 | 18 |
| Plateau | 22 | 22 | 25 |
| Basin | 28 | 27 | 22 |
| PCE (log) | 18 | 15 | 10 |
| GDP (log) | 2 | 3 | 2 |
| Population (log) | 4 | 4 | 4 |
| Married | 30 | 30 | 23 |
| *Region* |  |  |  |
| Yangtze River Delta | 25 | 29 | 14 |
| South East | 13 | 8 | 16 |
| Central | 23 | 32 | 21 |
| South West | 8 | 16 | 13 |
| North West | 24 | 31 | 20 |
| North East | 27 | 19 | 30 |
| Schooling | 3 | 2 | 3 |
| Social Participation | 29 | 24 | 28 |
| *Hukou* | 7 | 13 | 5 |
| Note: random forest model (RF) with mean decrease in accuracy criterion (MDA) | | | |

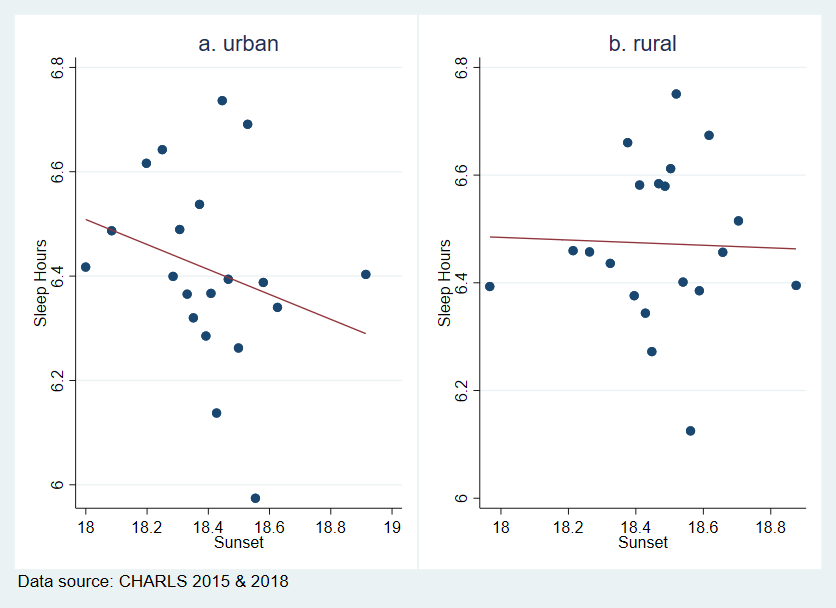

**Supplementary Figure 1** Binned scatterplots of sleep duration and sunset by *hukou* (non-agricultural employed)

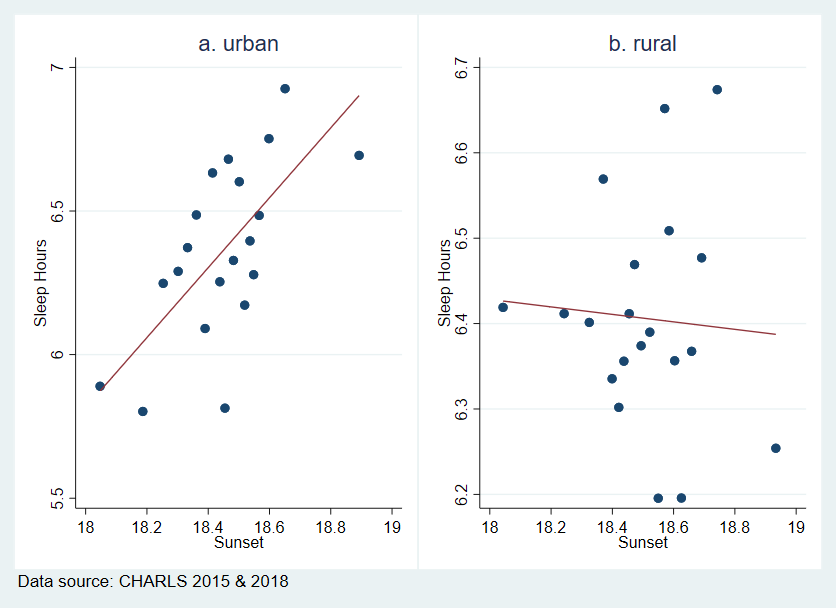

**Supplementary Figure 2** Binned scatterplots of sleep duration and sunset by *hukou* (self-employed)

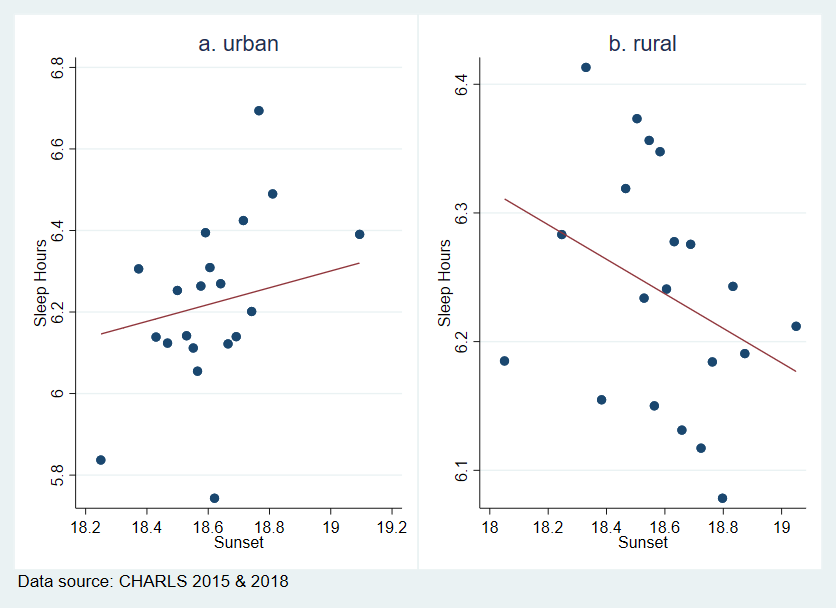

**Supplementary Figure 3** Binned scatterplots of sleep duration and sunset by *hukou* (farmer)

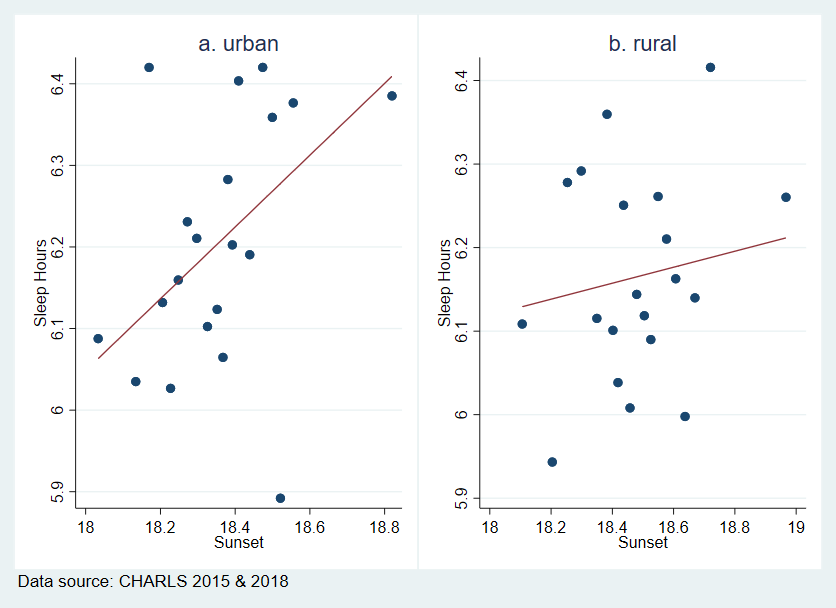

**Supplementary Figure 4** Binned scatterplots of sleep duration and sunset by *hukou* (no work)

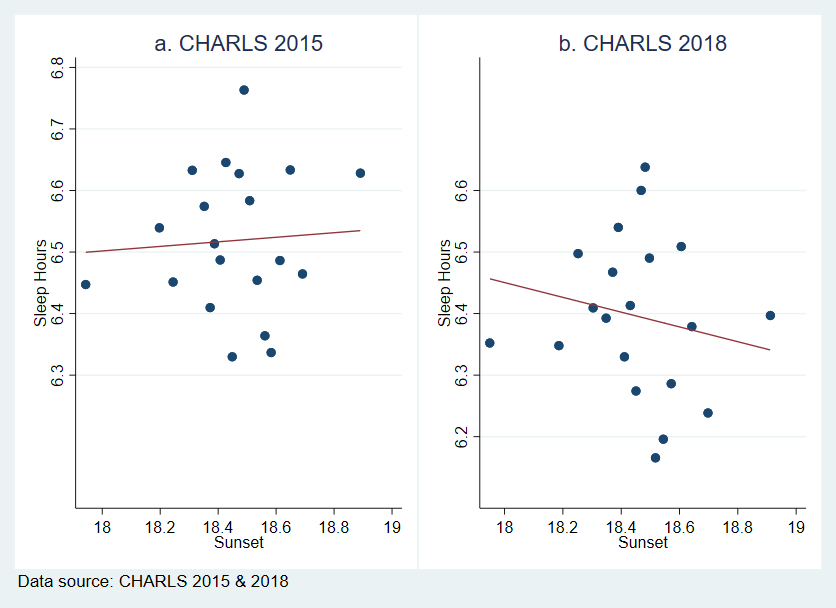

**Supplementary Figure 5** Binned scatterplots of sleep duration and sunset by wave (non-agricultural employed)

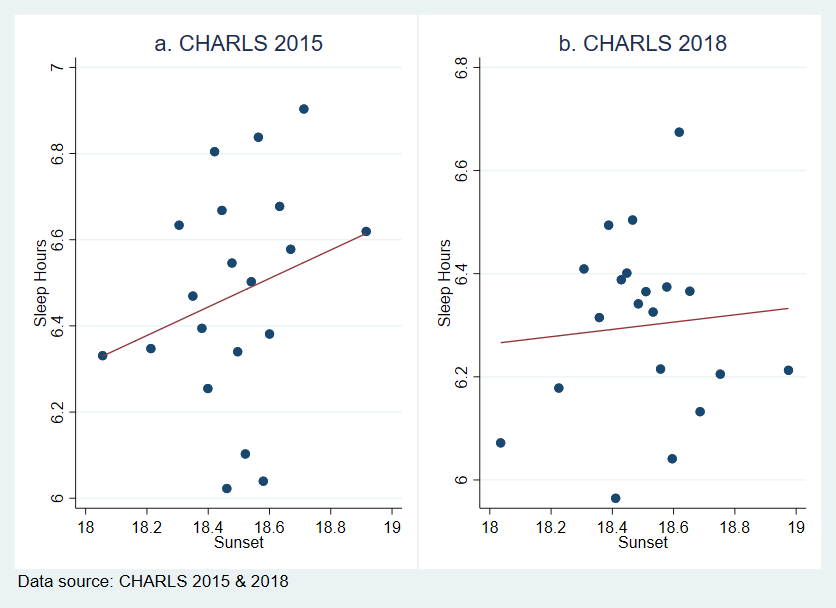

**Supplementary Figure 6** Binned scatterplots of sleep duration and sunset by wave (self-employed)

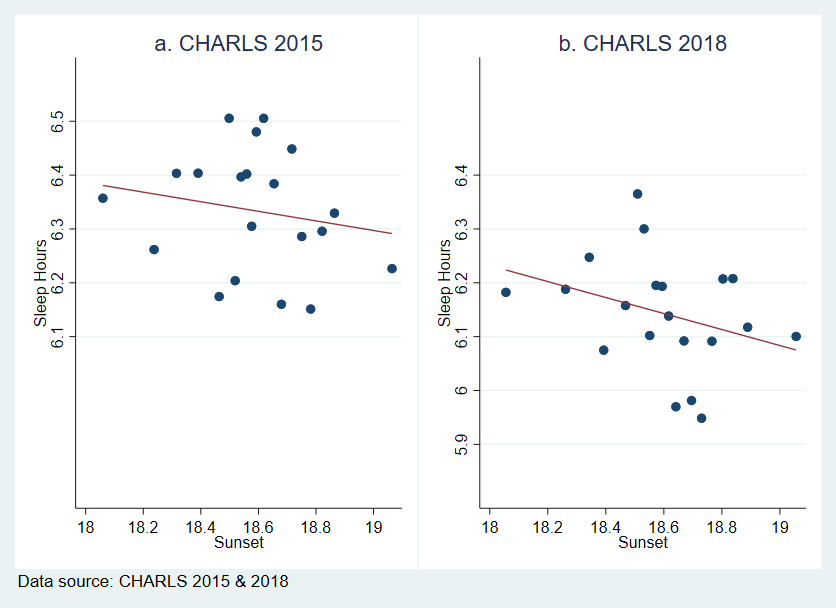

**Supplementary Figure 7** Binned scatterplots of sleep duration and sunset by wave (farmer)

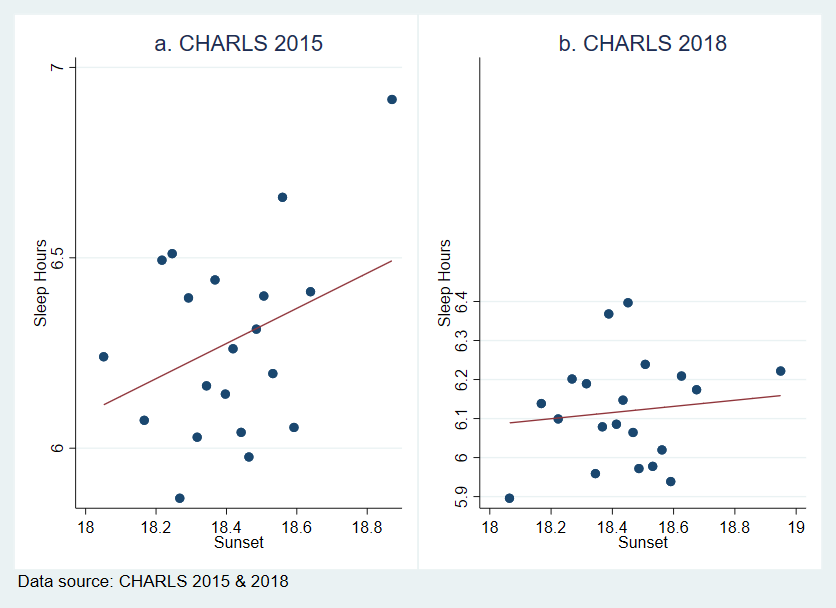

**Supplementary Figure 8** Binned scatterplots of sleep duration and sunset by wave (no work)
